# The use of angiotensin-converting enzyme inhibitors in hospitalized patients with COVID-19 is associated with a lower risk of mortality

**DOI:** 10.1101/2025.03.12.25323615

**Authors:** Mykola Khalangot, Vitaliy Gurianov, Tamara Zakharchenko, Victor Kravchenko, Olena Kovzun, Volodymyr Shupachynskiy, Mykola Tronko

**Author notes:** Corresponding author: Mykola Khalangot Komisarenko Institute of Endocrinology and Metabolism, 69 Vyshgorodska Str., Kyiv, 04114 Ukraine.

## Abstract

The relationship between the use of angiotensin-converting enzyme inhibitors (ACEIs) and angiotensin II receptor blockers (ARBs), diabetes mellitus (DM), and the risk of death in patients with COVID-19 remains controversial. We hypothesized that DM and certain characteristics of the COVID-19 course during hospital treatment may alter the assessment of the effect of ACEIs/ARBs on COVID-19 outcomes. The records of 153 COVID-19 inpatients admitted to a municipal clinic in Kyiv, Ukraine, between October and December 2021 were reviewed. To assess the effect of ARBs/ACEIs and other hypotensive drugs, a no hypotensives group was used for comparison. A multivariable logistic regression model was employed to assess the odds ratio (OR) of death. If DM was known at the time of hospitalization (n=28), there was a higher proportion of deaths compared to the group without DM (n=125): 53.6% vs. 12.8%, p < 0.001. After adjusting for age, minimal O2 saturation, DM, and antihypertensive therapy, the ACEIs-associated OR was 0.10 (0.02–0.69). The DM-associated OR was 8.25 (1.92–35.42). The use of ACEIs in the treatment of COVID-19 inpatients is associated with a lower risk of mortality compared to those not using hypotensive treatment, regardless of the presence of DM.

## Introduction

The end of the SARS-CoV-2 pandemic has not ended the state of uncertainty regarding the effect of angiotensin-converting enzyme inhibitors (ACEIs) and angiotensin II receptor blockers (ARBs) on the risk of mortality in patients with COVID-19: an observational study conducted in China [1] based on data from several hospitals found that ARB use was associated with lower all-cause mortality (adjusted HR, 0.53; 95% CI, 0.38-0.73; *p* < 0.001), while ACEI treatment did not affect mortality. However, in a similar study in Japan, ARBs did not significantly reduce in-hospital mortality over ACEIs in patients admitted for COVID-19, (adjusted OR, 0.95; 95% CI 0.69–1.3) [2]. Furthermore, the conclusions drawn by different reviews are markedly diverse. Xu J. et al. [3] assert that “there has been no evidence for initiating an ACEI/ARB regimen as prevention or treatment of COVID-19.” Conversely, Baral R. et al. [4], in their systematic review and meta-analysis of 52 studies involving 101,949 patients with COVID-19, identified a significantly lower risk of multivariable-adjusted mortality and severe adverse events among patients who received ACEIs or ARBs compared to those who did not. These authors suggest that ACEIs and ARBs may offer protective benefits, advocating for future randomized clinical trials to establish causality. It is notable, however, that the study by Baral R. et al. did not differentiate between the effects of ACEIs and ARBs. It is also concerning that in the meta-analysis by Xu J. et al., the criteria for excluding 4,415 out of 4,969 records were not clearly explained.

To date, the only RCT [5] conducted in Indian and Australian patients with mild COVID-19 who were randomly assigned to receive a low-dose ARB (40 mg/day telmisartan) failed to demonstrate a benefit of experimental treatment for COVID-19. The impact of diabetes both on the effects of ACEIs/ARBs in the context of COVID-19 and on COVID-19 outcomes unrelated to ACEIs/ARBs remains unclear.

Study in Ukraine found a very significant reduction in the risk of death for hospitalized patients with COVID-19 without DM, who received ACEIs/ARBs compared with the corresponding risk of normotensive COVID-19 inpatients who did not receive antihypertensive treatment: OR 0.22 (95% CI 0.07–0.72) adjusted for age, gender and fasting plasma glucose [6]. A recent retrospective multicentre European study [7] revealed no association between mortality and renin-angiotensin-aldosterone system inhibitor therapy in adults with diabetes admitted to hospital with COVID-19. In the last 2 studies, there was no separate assessment of the effects of ACEIs and ARBs on the outcomes of COVID-19.

The assessment of the relationship between DM and the risk of death in patients with COVID-19 is still controversial. On the one hand, a large population study in England confirmed the presence of such positive relationship: the odds ratios (ORs) for in-hospital COVID-19-related death were 3·51 (95% CI 3·16–3·90) in people with type 1 diabetes and 2·03 (1·97–2·09) in people with type 2 diabetes [8]. At the same time, another single-center study in England found no difference in mortalities based on the diabetes status [9], a similar result was reported by the authors in Morocco [10] and Ghana [11]. There is still no obvious explanation for the controversial effects of DM on mortality from COVID-19. We suggest that it is advisable to change the approaches to assessing the severity of patients, which may change during hospital treatment. This may refer to oxygen saturation, glycemia, and other indicators.

We also hypothesized that such approaches to characterization of the course of the disease may change the assessment of the effect of ACEI/ARBs on the outcomes of COVID-19 in patients with DM.

## Methods

### Study design and setting

The study was conducted by analyzing the archives of one of the multidisciplinary city hospital’s: Municipal Clinic – (MC), Kyiv, Ukraine. From October 2021 patients with clinical symptoms similar to Covid-19 were hospitalized in cardiologic and intensive care departments of this Hospital. Diagnosis of Covid-19, selection of patients for hospitalization, clinical examinations and treatment were performed according to relevant national standards [12], which were updated according to WHO recommendations. Detailed criteria for hospitalization were presented earlier [6].

A retrospective, observational study was carried out at the MC. The medical files of 153 patients, with a definite in-hospital outcome, admitted at MC between the periods of October 1 and December 31, 2021 were consecutively reviewed and the relevant data extracted.

### Patients characteristics

The diagnosis of Covid-19 was confirmed by polymerase chain reaction (PCR test, n = 145) or by detection of specific antibodies (IgM, n = 7), or specific antigen (express method, n=1). One patient with severe symptoms of COVID was included in the analysis despite lack of laboratory confirmation of SARS-CoV-2. Information on some clinical and anthropometric characteristics of patients: fasting plasma glucose (FPG), peripheral capillary oxygen saturation (SpO2); c-reactive protein (CRP), procalcitonin, D-dimer, sodium levels, body mass index (BMI); systolic and diastolic blood pressure (BP) level was extracted from patients’ paper documents.

The peculiarity of this study should be considered that we used not only the initial levels of some clinical and laboratory characteristics but tried to present their most clinically significant levels. Therefore, the highest (maximum) levels of BP, body temperature, CRP, D-dimer, Procalcitonin and the lowest (minimum) levels of SpO2 recorded during the hospital stay are presented. Regarding FPG and Sodium, we present both their maximum and minimum levels. This also applies to glucose-adjusted [13] sodium levels. Each patient’s treatment data were extracted from the paper files by two members of our research team and then checked and confirmed by a third person.

Similar to our recent study of the association between the use of certain antihypertensive drugs and COVID-19 outcomes [6] performed in Donetsk region of Ukraine, we categorized only previously known cases as DM. When evaluating COVID-19 outcomes depending on the use of certain types of hypotensive treatment, we used the results of patients who did not need hypotensive treatment as a reference group. However, in Kyiv study we were able to present more detailed treatment categories, i.e., we separated the results of ACEIs treatment from the results of ARBs.

Leaving the hospital due to recovery or death was considered as a dichotomous treatment outcome. 10 patients were transferred from MC to other city hospitals, their outcomes were also identified and taken into account in this study.

Statistical analysis has been performed using MedCalc® Statistical Software version 22.009 (MedCalc Software Ltd, Ostend, Belgium; https://www.medcalc.org; 2023). For the quantitative data mean and standard deviation (±SD) for normal distribution or median (Me) and interquartile range (QI – QIII) for non-normal distribution were presented. Comparisons of two groups were performed by t-test for normal distribution or Mann–Whitney test for non-normal distribution. For qualitative data the proportion (%) was presented, comparisons of two groups were performed by Fisher’s exact test. For the analysis association of the dependent variable with risk factors logistic regression analysis was used. To estimate the diagnostic efficiency of the logistic regression models area under the ROC curve (AUC) and its 95% CI were calculated. To estimate the effect of risk factors on the dependent variable the odds ratios (OR) and its 95% CI were calculated. In all of the tests the *p* value < 0.05 was considered significant.

## Results

In a group of patients with COVID-19, where DM was known at the time of hospitalization (n=28, table 1), there was a significantly higher proportion of deaths than in the group without information on the presence of DM (n=125): 53.6% vs. 12.8%, *p* <0.001.

**Table 1.** Anthropometric and laboratory characteristics and outcomes of hospitalized patients with COVID-19 depending on the presence of a history of diabetes mellitus (DM).

| Variables | Without DM<br>(n=125) | DM<br>(n=28) | *p* |
| --- | --- | --- | --- |
| Age, yrs | 70<br>(61.5 – 76.45) | 73.25<br>(67.250 – 81.15) | 0.079 |
| women, n (%) | 79 (63.2) | 17 (60.7) | 0.831 |
| Height, cm | 165<br>(162.250 – 170) | 165<br>(162.250 – 175) | 0.659 |
| Weight, kg | 80<br>(70 – 90) | 82.5<br>(72 – 96.5) | 0.279 |
| BMI, kg/m<sup>2</sup> | 29.1<br>(25.325 – 31.6) | 28.4<br>(26.550 – 34.6) | 0.341 |
| Systolic BP, maximal, mmHg | 140<br>(130 – 160) | 170<br>(140 – 180) | **0.003** |
| Diastolic BP, mmHg | 90 (80 – 90) | 90 (80 – 107.5) | 0.076 |
| Myocardial infarction<br>history, n (%) | 13 (11.0) | 5 (19.2) | 0.322 |
| Stroke history, n (%) | 7 (6.0) | 0 (0.0) | 0.347 |
| O<sub>2</sub> saturation, minimal, % | 88<br>(79.5 – 92) | 78<br>(65.5 – 90.5) | **0.006** |
| Temperature, maximal, °C | 38.09 (0.89) | 38.11 (0.82) | 0.882 |
| FPG maximal, mmol/L | 6.8 | 15.8 | **<0.001** |
|  | (5.8 – 8.525) | (11.6 – 22.850) |  |
| FPG minimal, mmol/L | 4.8 (3.9 – 5.7) | 4.7 (3.9 – 6.750) | 0.29 |
| CRP, maximal, mg/L | 50.2<br>(11.650 – 106) | 110.3<br>(49.750 – 126.250) | **0.009** |
| Procalcitonin, maximal,<br>ng/ml | 0.1<br>(0.1 – 0.2) | 0.23<br>(0.075 – 0.525) | 0.227 |
| D-Dimer, maximal, mg/L | 0.64<br>(0.320 – 1.450) | 0.84<br>(0.750 – 3.907) | **0.047** |
| Sodium minimal, mmol/L | 141<br>(139 – 144) | 141.5<br>(137.5 – 142) | 0.256 |
| Sodium maximal, mmol/L | 143<br>(141 – 145) | 143.5<br>(142 – 145.5) | 0.175 |
| Sodium minimal, corrected<br>mmol/L | 141<br>(139 – 143) | 141<br>(138 – 142) | 0.3 |
| Sodium maximal corrected,<br>mmol/L | 144<br>(141 – 146) | 148<br>(145.5 – 152) | **<0.001** |
| Duration of treatment, days | 13<br>(9 – 18) | 15<br>(10 – 20) | 0.471 |
| In hospital mortality, n (%) | 16 (12.8) | 15 (53.6) | **<0.001** |

Notes:
Data are presented by medians and inter quartile ranges (Q<sub>I</sub>-Q<sub>III</sub>) or ± SD. The comparison was based on the Mann-Whitney test or t-test. For qualitative variables frequency are presented, Fisher's exact test was used.

Both groups were equally dominated by women. A lower level of minimal O2 saturation, a higher level of maximal Systolic BP, FPG, CRP, D-Dimer and Sodium corrected were noted in the DM group. These groups did not differ in terms of gender, BMI, myocardial infarction, stroke history, or maximum body temperature. The median ages in the group without DM and with DM did not differ significantly (*p* = 0.079) and were equal to 70.0 and 73.25 years, respectively.

The treatment of patients in the groups without a history and with a history of diabetes did not differ with regard to the use of corticosteroids and antibiotics, while the use and structure of antihypertensive medications differed significantly between these groups (Table 2). More than a third of patients without a history of DM did not need treatment for arterial hypertension, while in the group with diabetes this situation applied to only one patient (3.6%). The proportion of ARB treatment was 15.8% and 32.1% for the non-DM and DM groups, respectively.

**Table 2.** Use of some types of treatment of COVID-19 inpatients depending on the history of diabetes.

| Variables |  | Without DM<br>n=125 (100) | DM<br>n=28 (100) | *p* |
| --- | --- | --- | --- | --- |
| Corticosteroids | Dexamethasone | 79 (63.2) | 17 (60.7) | 0.388 |
|  | No steroids | 13 (10.4) | 1 (3.6) |  |
|  | Methylprednisolone | 33 (26.4) | 10 (35.7) |  |
| Antibiotics | Intravenously | 86 (72.9) | 19 (70.4) | 0.741 |
|  | No antibiotics | 24 (20.3) | 5 (18.5) |  |
|  | Orally | 8 (6.8) | 3 (11.1) |  |
| Insulin | No | 120 (96.0) | 14 (50.0) | **<0.001** |
|  | Yes | 5 (4.0) | 14 (50.0) |  |
| Antihypertensives | No antihypertensives | 39 (32.5) | 1 (3.6) | **0.009** |
|  | ARB's | 19 (15.8) | 9 (32.1) |  |
|  | ACEi's | 37 (30.8) | 9 (32.1) |  |
|  | Other's | 25 (20.8) | 9 (32.1) |  |
| Other antidiabetes<br>treatments | No | 121 (96.8) | 15 (53.6) | **<0.001** |
|  | Yes | 4 (3.2) | 13 (46.4) |  |
| Mechanical<br>Ventilation (any type) | No | 112 (89.6) | 18 (64.3) | **0.002** |
|  | Yes | 13 (10.4) | 10 (35.7) |  |
| Invasive Mechanical<br>Ventilation | No | 114 (91.2) | 18 (64.3) | **0.001** |
|  | Yes | 11 (8.8) | 10 (35.7) |  |
| Noninvasive Mechanical Ventilation | No | 119 (95.2) | 22 (78.6) | **0.009** |
|  | Yes | 6 (4.8) | 6 (21.4) |  |

Note: percentages are given in parentheses.

A small proportion of patients without a history of DM were treated with insulin (4%) or other antidiabetic agents because of hyperglycemia detected during treatment for COVID-19. The use of mechanical lung ventilation in patients with DM occurred three times more often (35.7%) than in the group without DM (10.4%): RR=3.4 (95% CI 1.7 – 7.0). A similar ratio in the groups of COVID-19 inpatients without diabetes and with diabetes occurred regarding the share of use of both invasive and non-invasive mechanical ventilation (Table 2).

Univariable logistic regression modeling (table 3) indicates a positive relationship between age, history of diabetes, blood pressure, BMI, maximal levels of FPG, Sodium, Sodium corrected, CRP, D-Dimer and the risk of dying for COVID-19 inpatients. Minimal O2 saturation negatively and expectedly associated with the risk of dying. Assessment of treatment effects reveals a positive association with the odds of dying only for insulin and other anti-diabetes treatments (Table 3).

**Table 3.** Risks of death of COVID-19 inpatients (n=153) estimated using univariable logistic regression models.

| Factors |  | Model coefficient<br>b±m | *p* | OR (95% CI) |
| --- | --- | --- | --- | --- |
| Age, yrs |  | 0.043±0.019 | **0.02** | **1.04 (1.00–1.08)** |
| Gender | women | Reference |  |  |
|  | men | 0.08±0.41 | 0.851 | – |
| Myocardial infarction history |  | -0.27±0.67 | 0.687 | – |
| Stroke history |  | 0.48±0.86 | 0.579 | – |
| Diabetes mellitus history |  | 2.06±0.46 | **<0.001** | **7.86 (3.17–19.52)** |
| BMI, kg/m<sup>2</sup> |  | 0.089±0.039 | **0.022** | **1.09 (1.01–1.18)** |
| Systolic BP maximal, mmHg |  | 0.033±0.009 | **<0.001** | **1.03 (1.02–1.05)** |
| Diastolic BP, mmHg |  | 0.036±0.016 | **0.024** | **1.04 (1.00–1.07)** |
| Temperature, maximal, °C |  | 0.23±0.23 | 0.327 | – |
| O2 Saturation minimal, % |  | -0.15±0.03 | **<0.001** | **0.86 (0.81–0.91)** |
| FPG maximal, mmol/L |  | 0.12±0.03 | **<0.001** | **1.12 (1.06–1.19)** |
| Procalcitonin maximal, ng/ml |  | 0.039±0.029 | 0.188 | 1.04 (0.98–1.10) |
| CRP maximal, mg/L |  | 0.0091±0.0045 | **0.046** | **1.01 (1.00–1.02)** |
| D-Dimer maximal, mg/L |  | 0.20±0.10 | **0.042** | **1.22 (1.01–1.49)** |
| Sodium minimal, mmol/L |  | -0.075±0.042 | 0.073 | 0.93 (0.86–1.01) |
| Sodium maximal, mmol/L |  | 0.12±0.05 | **0.029** | **1.12 (1.01–1.25)** |
| Sodium minimal, corrected mmol/L |  | -0.064±0.041 | 0.116 | – |
| Sodium maximal corrected, mmol/L |  | 0.17±0.05 | **<0.001** | **1.19 (1.087–1.30)** |
| Insulin |  | 1.79±0.52 | **0.001** | **5.98 (2.17–16.48)** |
| Other anti-diabetes treatments |  | 1.76±0.54 | **0.001** | **5.83 (2.03–16.76)** |
| Corticosteroids |  | -19.5±6300 | >0.05 | – |
| Antibiotics | Intravenously | Reference |  |  |
|  | None | -1.39±0.77 | 0.069 | 0.25 (0.06–1.12) |
|  | Orally | 0.23±0.71 | 0.750 | – |
| Antihypertensives | None | Reference |  |  |
|  | ACEIs | -0.72±0.60 | 0.234 | – |
|  | Other's | 0.36±0.54 | 0.498 | – |
|  | ARB's | 0.47±0.56 | 0.402 | – |

Multivariate logistic regression analysis was performed for evaluation the risk of death. A stepwise method was used (entering a variable if *p*<0.05 and removing a variable if *p*>0.1). Selected significant characteristics in the multivariate model: Age, Antihypertensives, O2 Saturation minimal, DM history (table 4).

**Table 4.** Analysis of a multivariable logistic regression model for predicting the risk of death for inpatients with COVID-19.

| Factors |  | Model coefficient<br>b±m | *p* | OR (95% CI) |
| --- | --- | --- | --- | --- |
| Age, yrs |  | 0.057±0.027 | **0.033** | **1.06 (1.00–1.12)** |
| O2 Saturation minimal, % |  | -0.17±0.03 | **<0.001** | **0.84 (0.79–0.90)** |
| Diabetes mellitus history |  | 2.11±0.74 | **0.005** | **8.25 (1.92–35.42)** |
| Antihypertensives | None | Reference |  |  |
|  | ACEIs | -2.26±0.97 | **0.019** | **0.10 (0.02–0.69)** |
|  | Other's | -0.17±0.78 | 0.826 | — |
|  | ARB's | -0.65±0.96 | 0.499 | — |

Area under ROC curve of the model AUC= 0.94 (95% CI 0.89 – 0.97) which indicates a strong association of selected variables with the risk of death. Regardless of age, O2 Saturation, and Diabetes mellitus history, treatment of inpatients with COVID-19 with ACEIs was associated with a significant reduction in the risk of death compared to no antihypertensive treatment: OR (95% CI) 0.10 (0.02–0.69). However, the estimated OR (95% CI) associated with diabetes mellitus history within this model was very high: 8.25 (1.92–35.4).

The addition of other variables the risk of which was significant when assessed using univariable logistic regression models (table 3) in the multivariable model led to the loss of their significance and did not lead to an improvement in the prognostic characteristics of the model, which indicates their association with the already applied variables.

Since two variables in this multivariate model were continuous and the other four categorical, we ran a similar model that included only categorical variables: age > 70 years and O2 saturation minimal ≤ 82% (cut-offs based on median level and better Joden index, respectively). This model demonstrated the same odds directions: higher mortality risk for DM and lower mortality risk for ACEIs (Figure S1). The AUC of this model was 0.937 (95% CI 0.86 - 0.97), which was very close to the previous model.

Additionally, we conducted an analysis using a multivariable logistic regression model to predict the risk of death exclusively for inpatients with COVID-19 who tested positive via PCR, adding the maximal systolic BP variable. Excluding several patients without PCR confirmation and adding this variable did not significantly alter the association between ACEI treatment and the risk of COVID-19-related death (Table S1). The area under the ROC curve of the model, AUC = 0.95 (95% CI 0.89 – 0.98), indicates a strong association of the selected variables with the risk of death.

## Discussion

In our relatively small (n=153) sample of older (median age 70 yrs) COVID-19 inpatients with a significant proportion of patients with DM history (18.3%), a rather large proportion of deaths (20.26%) was found. Recent studies from China report mortality in an age-matched population of 5.2% and 8.6% (ARB- and ACEI -treated COVID-19 inpatients respectively) [1], similar data from Japan were 5.2% and 7.7% [2]. However, in 2020, one of the hospitals in England reported a mortality rate of 38.4% for all hospitalized patients with COVID-19 (n=232), and 46% for patients with diabetes [9]. Most likely, these discrepancies indicate different criteria or practice of their application for hospitalization of COVID-19 in different countries or clinical centers.

Our analysis revealed that the majority (53.6%) of hospitalized patients with DM history unfortunately died, while for patients without pre-existing DM the mortality was 12.8%. Diabetes-related risk of death estimated using a one-way logistic regression model, OR (95% CI) was 7.86 (3.17–19.52). A similar regression analysis indicated a statistically significant mortality risk associated with increasing age and BMI (table 3), that is, it is impossible not to consider at least these factors when assessing the contribution of DM to the mortality risk of patients with COVID-19. To a large extent, this may also apply to systolic and diastolic maximal BP and indicators reflecting the course of DM and COVID-19 (minimal SpO2, maximum levels of FPG, Sodium, Sodium corrected, CRP, D-Dimer) that have revealed a connection with the risk of death in patients with COVID-19. Thus, when evaluating COVID-19 death risks, we evaluated SpO2, FPG, sodium and some cytokines like minimal and/or maximal levels, instead of their levels at the time of admission, which was used by most other studies. Regarding FPG (but not SpO2), an approach similar to ours was used by Italian researchers [14]. According to our assessment, the higher the minimum SpO2 level for a particular patient, the lower the risk of dying, which is the expected result (table 3).

Logistic regression analysis was applied to evaluate the effects of the independent variables on the risk of death. Stepwise method was used to find the best fitting model which described the relationship between the risk of death and a set of independent variables in a multifactor logistic regression model.

As a result of this analysis, we can conclude that regardless of age, minimum SpO2 and history of DM, treatment of inpatients with COVID-19 with ACE inhibitors was associated with a significant reduction in the risk of death compared to no antihypertensive treatment: OR (95% CI) 0.10 (0.02–0.69). However, the estimated risk associated with a history of diabetes in this model was very high: OR (95% CI) 8.25 (1.92–35.42). It should be noted that due to the use of such a statistical model, we were able to show a relationship between the use of ACEIs and a decrease in the chances of dying for patients with COVID-19, while this relationship was not detected during univariate analysis (table 3). To our knowledge, a similar analytical approach has not previously been applied to assess COVID-19 outcomes.

Researchers from Morocco reported an increase in CRP and D-Dimer levels in COVID-19 inpatients with DM, but unlike us, they did not find a significant difference in the mortality of COVID-19 inpatients depending on DM. To clarify the relationship between COVID-19 mortality and DM, they suggest investigating the interaction of medications with ACE-2 receptors (that is, the use of ARBs and ACEIs in treatment) and the role of co-morbidities [10]. Resent multivariable logistic regression mortality analysis of COVID-19 patients with type 2 diabetes mellitus in Istanbul (Turkey) confirmed an independent mortality risk related with increasing age, male gender, obesity, lymphopenia and insulin treatment, but did not reveal such a relationship with ARBs / ACEIs treatment, high HbA1c and some comorbidities [15]. It should be noted that the study in Istanbul concerned the entire population, and not only those patients who, due to the severity of their condition, required hospitalization. A large retrospective study in China has just presented an analysis of adjusted hazard ratios for risk factors associated with in-hospital all-cause mortality in the multivariate Cox regression model. According to this model, treatment of hypertension with ARBs (but not ACEIs) was found to be associated with a significant reduction in the risk of death for patients hospitalized for COVID-19 [1]: “After adjusting for age, gender, comorbidities, and in-hospital medications (such as metformin and statins), the use of ARBs was associated with lower all-cause mortality (adjusted HR 0.54; 95% CI, 0.39–0.74; *p* < 0.001). Besides, the adjusted HRs for association of other variables for all-cause mortality were 1.91 (95% CI, 1.63–2.23), 1.53 (95% CI, 1.35–1.75), 0.59 (95% CI, 0.47–0.73), 0.51(95% CI, 0.31–0.85), and 1.63 (95% CI, 1.43–1.86) for age, gender, statins, metformin, and hypercholesterolemia, respectively”. In our opinion, an interesting result of this study, which its authors for some reason did not pay enough attention to, is the assessment of DM and its treatment with metformin as a protective factor for mortality in COVID-19 inpatients, while insulin treatment is associated with a significant increase in the risk of death in these patients. These results are consistent with our data on the risk associated with the treatment of patients with type 2 diabetes with insulin: OR 5.98 (2.17–16.48) using one-way logistic regression model. Risk connected with DM history: OR 10.14 (3.45–29.83) using a multifactor (age and antihypertensive treatment) logistic regression model for predicting the risk of death for inpatients with COVID-19.

If the high insulin-related risk of mortality in patients with COVID-19 is confirmed by studies from England [16] and China [17], the protective effect of DM shown by authors from China [1] is not at all an expected result. A study from England is rather coincident with our inpatients-based study: the odds ratios (ORs) for in-hospital COVID-19-related death were 2.03 (1.97– 2.09) in people with type 2 diabetes [8]. Using a multivariable logistic regression model (age, SpO2, DM history, stratified antihypertensive therapy) to predict the risk of death for hospitalized patients with COVID-19 found ACEI-associated ORs of 0.10 (0.02–0.69) or 0.18 (0.04–0.70) depending on SpO2 on/off.

Our estimate of the association of treatment with ARBs or ACEIs with the risk of death in inpatients with COVID-19 does not agree with corresponding estimates recently made in Japan [2] and China [1]. Using a logistic model of analysis, Japanese authors found that among hypertensive patients hospitalized for COVID-19, ARB was associated with lower crude rate of in-hospital mortality, but not in multivariate-adjusted analysis [2]. The described sample of data from Japan is unique in that there is no use of non-invasive mechanical ventilation and no data on antibiotics. The proportion of deaths was at least 2 times lower than in the Ukrainian sample of hospitalized patients, which may indicate less strict criteria for hospitalization and partially explain the different results of the studies. A study from China [1] reported almost the same mortality of hospitalized patients as in Japan, but ARB administration was significantly associated with a lower risk of all-cause mortality in patients with COVID-19. This conclusion was made by multivariate Cox regression analysed and Kaplan–Meier curves. The use of Cox regression in this case may have flaws that could affect the result. The use of Cox proportional-hazards regression method to analyze the influence of factorial features on mortality risk, in this case may lead to estimate bias (the best method for analyzing these data would be using logistical regression models).

Italian researchers (Piarulli, F., et al. 2023) reported mortality among 389 COVID-19 patients with diabetes hospitalized in the early period of the pandemic in the Veneto region: 22.89% [14]. Using multivariable logistic regression analysis, these authors reported that previous use of RAAS-i was associated with both reduced hospitalization and best outcomes in the diabetic population hospitalized for COVID-19. This finding is largely consistent with our results, but we were able to conduct separate analyzes for ARBs and ACEIs. In addition, we used a stepwise method to find the best fitting in a multifactor logistic regression model. Besides, we analyzed the relationship of treatment outcomes with the actual intake of medication during hospitalization, but not with the treatment that preceded it.

One of the features of our approach is that the effect of using ARBs / ACEIs and other hypotensive drugs was not compared between these groups, but the results of “No hypotensives” were used as a comparison group. We used a similar approach earlier when analyzing the results of hospital treatment of COVID-19 (n=1097) depending on the use of RAAS inhibitors (ARBs / ACEIs). We found a reduction in the risk of death for COVID-19 inpatients without DM, who received RAAS inhibitors compared with the corresponding risk of normotensive inpatients, who did not receive antihypertensives: OR 0.22 (95% CI 0.07–0.72) adjusted for age, gender and FPG. The mentioned study did not distinguish the effect of ARBs from the effect of ACEIs and the protective effect was shown only for patients without DM [6].

A small sample size can be considered as a weakness of our study. However, large sample size in the studies of Xu K, et al., 2023 [1] and Hamada, S., et al., 2023 [2] did not prevent them from obtaining conflicting results. A possible explanation may be that in the case of working with large medical databases and not with paper medical documents, often only the medications that patients took before hospitalization are considered. In our study, only actual antihypertensive treatment in the hospital was compared. In addition, we believe that the small sample size may be a source of doubt rather in the case of reporting a negative result, while in our case the null hypothesis is rejected with statistically sufficient probability.

We emphasize that one of the strengths of our study is the investigation of the effects of ACEIs/ARBs in the Ukrainian population of patients with severe COVID-19. In a recent review, Sodhi P. et al., 2023 [18], highlight the role of ACE2 as a regulator of the RAAS and as a receptor for SARS-CoV-2. They discuss the significance of the two forms of ACE2, mACE2 and sACE2, and suggest that the interplay between SARS-CoV-2, mACE2, sACE2, and ANG II warrants further investigation in relation to COVID-19 severity. Additionally, they point out that ACE2 variants, which have been identified as specific to different populations, highlight interindividual variability.

A recently published opinion (Tsukamoto S. 2024) [19] explains the mechanism, due to which ACEIs have advantages over ARBs in the prevention and treatment of viral pneumonias: “One mechanism of ACE-I prevention of pneumonia is speculated to be an increase in the cough reflex. Bradykinin and substance P, which are normally metabolized by angiotensin-converting enzyme, accumulate in the airways after ACE-I administration, stimulating vagal afferent fibers that control the cough reflex, thereby promoting the cough reflex. Increased coughing may promote expectoration of pathogens, including microaspiration, and may contribute to the prevention of pneumonia. This mechanism is not present in angiotensin II receptor blockers (ARBs), which are also renin-angiotensin system (RAS) inhibitors”. Fastbom, J., et al. & Nyman Iliadou, A., 2024 conducted a cohort study, based on record linked individual-based data from national registers, of all Swedish inhabitants 50 years and older (n = 3,909,321) at the start of the first SARS-CoV-2 wave in Sweden. Lower risk of dying in COVID-19 was observed for ACEIs: HR 0.92 (0.86-0.99). For ARBs no significant associations was found [20].

Thus, our results probably received a theoretical explanation [19] and partial confirmation at a large population level [20]. We highlight the distinct aspects of our research, which include: The analyzed population of patients with a very severe course of COVID-19 and high mortality, especially among those with a history of diabetes mellitus (DM).

The identification of a DM-independent association between ACEI treatment during hospitalization and COVID-19 outcomes.

The results of the research that we present now are the basis for the following conclusion: regardless of age, minimal O2 saturation, DM history, treatment of inpatients with COVID-19 with ACEIs was associated with a significant reduction in the risk of death compared to no antihypertensive treatment.

## Data Availability

All data produced in the present study are available upon reasonable request to the authors

## Declarations

### Ethics approval and consent to participate

This study was conducted in accordance with the principles of the Declaration of Helsinki and approved by the Institution ethics committee. Owing to the retrospective nature and anonymity of this study, the ethics committee waived the need for written informed consent. Protocol # 35/4-KE from 01/04/2021.

### Consent for publication

Not applicable.

### Funding

This study was supported by the National Research Foundation of Ukraine **Grant# 2021.01/0213** “Study of the course and consequences of COVID-19 in patients with diabetes mellitus and the impact of SARS-CoV-2 infection on the rate of biological aging“

### Competing interests

The authors declared no potential conflicts of interest with respect to the research, authorship, and/or publication of this article.

### Availability of Data and Materials

The datasets generated during the current study are not publicly available, but de-identified data will be made available upon request where such requests are compliant with receipt of ethical approval from the sending and receiving hosts’ Institutional Ethics Review Boards.

## Acknowledgements

All authors are grateful to Ms. Oksana Opanasenko for technical assistance in conducting this study.

## Notes

### Competing Interest Statement

The authors have declared no competing interest.

### Author Declarations

This study was approved by the Komisarenko Institute of Endocrinology and Metabolism ethics committee. Owing to the retrospective nature and anonymity of this study, the ethics committee waived the need for written informed consent. Protocol # 35/4-KE from 01/04/2021.

## References

1. Xu K, He W, Yu B, Zhong K, Zhou D, Wang DW. Beneficial Effects of Angiotensin II Receptor Blockers on Mortality in Patients with COVID-19: A Retrospective Study from 2019 to 2020 in China. Cardiovasc Drugs Ther. 2023 Aug 11. doi: 10.1007/s10557-023-07494-5.

2. Hamada, S., Suzuki, T., Tokuda, Y., Taniguchi, K. and Shibuya, K., 2023. Comparing clinical outcomes of ARB and ACEi in patients hospitalized for acute COVID-19. Scientific Reports, 13(1), p.11810.

3. Xu, J., Teng, Y., Shang, L., Gu, X., Fan, G., Chen, Y., Tian, R., Zhang, S. and Cao, B., 2021. The effect of prior angiotensin-converting enzyme inhibitor and angiotensin receptor blocker treatment on coronavirus disease 2019 (COVID-19) susceptibility and outcome: a systematic review and meta-analysis. Clinical Infectious Diseases, 72(11), pp.e901–e913.

4. Baral R, Tsampasian V, Debski M, et al. Association Between Renin-Angiotensin-Aldosterone System Inhibitors and Clinical Outcomes in Patients With COVID-19: A Systematic Review and Meta-analysis. JAMA Netw Open. 2021;4(3):e213594. doi:10.1001/jamanetworkopen.2021.3594

5. Jardine, M.J., Kotwal, S.S., Bassi, A., Hockham, C., Jones, M., Wilcox, A., Pollock, C., Burrell, L.M., McGree, J., Rathore, V. and Jenkins, C.R., 2022. Angiotensin receptor blockers for the treatment of covid-19: pragmatic, adaptive, multicentre, phase 3, randomised controlled trial. BMJ 2022;379:e072175

6. Khalangot M, Sheichenko N, Gurianov V, Zakharchenko T, Kravchenko V, Tronko M. RAAS inhibitors are associated with a better chance of surviving of inpatients with Covid-19 without a diagnosis of diabetes mellitus, compared with similar patients who did not require antihypertensive therapy or were treated with other antihypertensives. Front Endocrinol (Lausanne). 2023 Jan 19;14:1077959.

7. Harris S, Ruan Y, Wild SH, Wargny M, Hadjadj S, Delasalle B, et al. Association of statin and/or renin-angiotensin-aldosterone system modulating therapy with mortality in adults with diabetes admitted to hospital with COVID-19: A retrospective multicentre European study. Diabetes Metab Syndrome: Clin Res Rev (2022) 16(5):102484. doi: 10.1016/j.dsx.2022.102484

8. Barron, E., Bakhai, C., Kar, P., Weaver, A., Bradley, D., Ismail, H., Knighton, P., Holman, N., Khunti, K., Sattar, N. and Wareham, N.J., 2020. Associations of type 1 and type 2 diabetes with COVID-19-related mortality in England: a whole-population study. The lancet Diabetes & endocrinology, 8(10), pp.813–822.

9. Alkundi A, Mahmoud I, Musa A, Naveed S, Alshawwaf M. Clinical characteristics and outcomes of COVID-19 hospitalized patients with diabetes in the United Kingdom: A retrospective single centre study. Diabetes Res Clin Pract. 2020 Jul;165:108263. doi: 10.1016/j.diabres.2020.108263.

10. Elamari, S., Motaib, I., Zbiri, S., Elaidaoui, K., Chadli, A. and Elkettani, C., 2020. Characteristics and outcomes of diabetic patients infected by the SARS-CoV-2. Pan African Medical Journal, 37:32.

11. Hardy, Y.O., Libhaber, E., Ofori, E., Amenuke, D.A.Y., Kontoh, S.A., Dankwah, J.A., LarsenCReindorf, R., OtuCAnsah, C., HuttonCMensah, K., Dadson, E. and Adamu, S., 2023. Clinical and laboratory profile and outcomes of hospitalized COVIDC19 patients with type 2 diabetes mellitus in Ghana–A singleCcenter study. Endocrinology, Diabetes & Metabolism, 6(1), p.e391.

12. STANDARDS OF MEDICAL CARE “CORONAVIRUS DISEASE (COVID-19)” ministry of health of Ukraine (Document in Ukrainian). Available at: https://moz.gov.ua/uploads/3/19713-standarti_med_dopomogi_covid_19.pdf.

13. Hillier TA, Abbott RD, Barrett EJ. Hyponatremia: evaluating the correction factor for hyperglycemia. Am J Med 1999;106:399–403.

14. Piarulli, F., Carollo, M., Ragazzi, E., Benacchio, L., Piovanello, F., Simoncello, I. and Lapolla, A., 2023. Association of COVID-19 outcomes with diabetes in the Veneto region (north-east italy): Epidemiological insights for the endemic phase?. Nutrition, metabolism, and cardiovascular diseases: NMCD, 33(11), pp.2141–2150.

15. Satman, I., Demirci, I., Haymana, C., Tasci, I., Salman, S., Ata, N., Dagdelen, S., Sahin, I., Emral, R., Cakal, E. and Atmaca, A., 2021. Unexpectedly lower mortality rates in COVID-19 patients with and without type 2 diabetes in Istanbul. diabetes research and clinical practice, 174, p.108753.

16. Khunti, K., Knighton, P., Zaccardi, F., Bakhai, C., Barron, E., Holman, N., Kar, P., Meace, C., Sattar, N., Sharp, S. and Wareham, N.J., 2021. Prescription of glucose-lowering therapies and risk of COVID-19 mortality in people with type 2 diabetes: a nationwide observational study in England. The lancet Diabetes & endocrinology, 9(5), pp.293–303.

17. Yu, B., Li, C., Sun, Y. and Wang, D.W., 2021. Insulin treatment is associated with increased mortality in patients with COVID-19 and type 2 diabetes. Cell metabolism, 33(1), pp.65–77.

18. Sodhi PV, Sidime F, Tarazona DD, Valdivia F, Levano KS. A Closer Look at ACE2 Signaling Pathway and Processing during COVID-19 Infection: Identifying Possible Targets. Vaccines. 2023; 11(1):13. 10.3390/vaccines11010013

19. Tsukamoto, S. Do angiotensin-converting enzyme inhibitors not reduce the risk of pneumonia?. Hypertens Res 47, 2961–2963 (2024). 10.1038/s41440-024-01848-8

20. Fastbom, J., Jonasdottir Bergman, G., Holm, J., Hanberger, H., Strålin, K., Walther, S., Alfredsson, J., State, M., Borg, N. and Nyman Iliadou, A. Use of drugs for hypertension or heart failure and the risk of death in COVID-19: association with loop-diuretics. Eur J Clin Pharmacol 80, 1515–1522 (2024). 10.1007/s00228-024-03709-2

