## Supplementary material for "The use of angiotensin-converting enzyme inhibitors in hospitalized patients with COVID-19 is associated with a lower risk of mortality": Table S1 Fig.S1

**Table S1.**  Analysis of a multivariable logistic regression model for predicting the risk of death for PCR test positive inpatients with COVID-19.

| Factors | | Model coefficient b±m | p | OR (95% CI) |
| --- | --- | --- | --- | --- |
| Age, yrs | | 0.052±0.032 | 0.098 | 1.05 (0.99–1.12) |
| Systolic BP, maximal, mmHg | | 0.029 ± 0.017 | 0.080 | 1.03 (0.99 – 1.06) |
| O2 Saturation minimal, % | | **-0.15±0.04** | **<0.001** | **0.86 (0.80–0.92)** |
| Diabetes mellitus history | | **1.96±0.80** | **0.014** | **7.11 (1.50–33.76)** |
| Antihypertensives | None | Reference | | |
|  | ACEi’s | **-2.24±1.06** | **0.035** | **0.11 (0.01–0.85)** |
|  | Other’s | 0.28±0.84 | 0.737 | – |
|  | ARB’s | -0.87±1.11 | 0.432 | – |

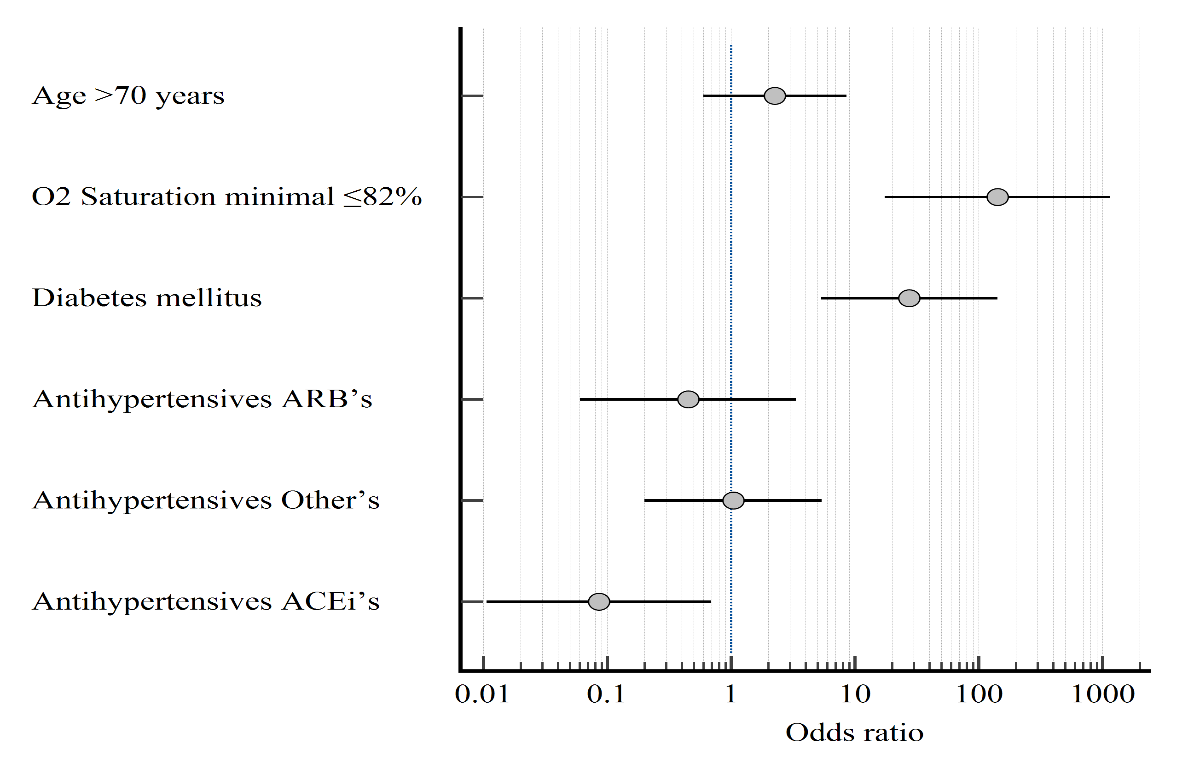

Figure S1. Odds Ratios with their 95% Confidence Intervals obtained from the analysis of a multivariable “categorical” logistic regression model for predicting the risk of death for inpatients with COVID-19.
